# Prediction of brucellosis incidence in China’s five highest-incidence provinces: Comparing time-series models with multi-source environmental predictors

**DOI:** 10.64898/2026.07.09.26357632

**Authors:** Yawen Qin, Qianran Gao, Haoran Liu, Hanjing Fan, Qingqing Wang, Wenyi Zhang, Changping Li, Qiulan Chen, Zhuang Cui

**Affiliations:** Department of Epidemiology and Biostatistics, School of Public Health, Tianjin Medical University, Tianjin, China; Chinese PLA Center for Disease Control and Prevention, Beijing, China; Department of Epidemiology, School of Public Health, China Medical University, Shenyang, China; National Key Laboratory of Intelligent Tracking and Forecasting for Infectious Disease, Chinese Center for Disease Control and Prevention, Beijing, China

**Keywords:** Brucellosis, Time-series forecasting, LSTM, SARIMAX, region-specific forecasting, Environmental variables

## Abstract

**Background:** Brucellosis is a severe zoonotic disease with pronounced seasonality and regional heterogeneity in high-incidence areas of China. Reliable forecasting tools are needed to inform prevention strategies, but the optimal modeling approach across different regions remains unclear.

**Principal Findings:** We collected monthly brucellosis incidence and 17 environmental variables from 2014 to 2024 across five high-incidence provinces: Inner Mongolia, Xinjiang, Shanxi, Heilongjiang, and Hebei. A three-step procedure—cross-correlation analysis, multicollinearity diagnostics, and stepwise regression—was used to select exogenous predictors. We then compared four time-series models: seasonal autoregressive integrated moving average (SARIMA), SARIMA with exogenous variables (SARIMAX), long short-term memory (LSTM), and LSTM with exogenous variables (LSTMX). All five provinces showed a unimodal seasonal pattern with peaks between April and July, though environmental drivers and optimal lag periods varied substantially by region, ranging from 1 to 6 months. In forecasting performance, LSTM achieved the highest accuracy in Shanxi (R²=0.925), Hebei (R²=0.876), and Xinjiang (R²=0.829), outperforming SARIMA and SARIMAX. LSTMX performed best in Inner Mongolia (R²=0.759) and Heilongjiang (R²=0.772) but showed weaker performance than LSTM in Shanxi and Hebei. Overall, adding exogenous variables did not consistently improve predictions across provinces.

**Conclusions:** Our findings demonstrate that LSTM-based models offer clear advantages for brucellosis forecasting in most high-incidence provinces, but the value of incorporating environmental predictors is region-dependent. These results support the development of tailored early warning systems and precision prevention strategies for brucellosis in high-risk areas of China.

**Author Summary:** Brucellosis is a bacterial disease that spreads from animals to humans, posing serious health risks, especially to livestock workers. Although preventable, its incidence has been rising in parts of China, highlighting the urgent need for more accurate forecasting tools. Traditional statistical models often fail to capture the complex patterns driving disease outbreaks, so we turned to deep learning. We developed an LSTM model—a form of artificial intelligence that learns from historical data—and compared its performance against standard models across five high-incidence provinces. Our LSTM model performed remarkably well, explaining up to 92.5% of the variation in brucellosis cases in some high-risk areas. Interestingly, adding environmental factors like temperature and humidity improved predictions only in certain provinces, not all. This finding underscores that no single model works everywhere; the best approach depends on local conditions. Our work advances the development of smarter, real-time early warning systems for brucellosis and other zoonotic diseases, aligning with the One Health approach that recognizes the interconnectedness of human, animal, and environmental health.

## 1. Background

Brucellosis is a zoonotic bacterial infection caused by *Brucella* species^1^, mainly infecting domestic animals such as cattle, sheep, and goats. The species most commonly associated with human disease are *B. melitensis*, *B. abortus*, *B. suis*, and *B. canis*^1,2^. Humans acquire infection through direct contact with infected animals, consumption of contaminated animal products (especially unpasteurized milk or cheese), or inhalation of infectious aerosols. Clinical presentation is nonspecific, including fever, sweats, fatigue, headache, and arthralgia, making it difficult to distinguish from other febrile illnesses^1,2^. Symptoms may persist for weeks to months, with complications such as endocarditis and neurological disorders^3^. Chronic brucellosis is defined as symptom persistence for more than twelve months^4,5^.

Brucellosis is among the most widespread zoonotic diseases worldwide. Recent estimates suggest that annual global incidence may reach 1.6–2.1 million cases, indicating that the disease burden has likely been underestimated^6^. The disease remains persistently endemic across Asia and Africa, with the highest incidence rates reported in Syria, Kyrgyzstan, Mongolia, Iran, Algeria, and Kenya. Asia bears the heaviest burden, particularly in East, Central, and West Asian countries. Over the past decade, the epidemiological range has expanded markedly, with affected countries increasing from 53 to at least 97^7^. Expansion of animal industries, urbanization, and inadequate hygienic measures in animal husbandry and food handling contribute to this sustained public health hazard^1^.

Brucellosis remains an urgent public health challenge in China. Nationally, human brucellosis incidence increased overall between 2005 and 2021^8^, declined during 2015–2018, and rebounded sharply after 2019^9^. Marked heterogeneity was observed across regions. Inner Mongolia bore the highest burden, with 87,961 cumulative cases and an average annual incidence of 71.88 per 100,000 population, ranking first nationwide for ten consecutive years. Xinjiang reported 27,845 cases, with the fastest growth among highest-incidence regions (average annual increase of 16.76%). Shanxi documented 21,932 cases (annual increase of 9.61%), Hebei reported 19,508 cases (annual increase of 4.63%), and Heilongjiang reported 17,637 cases (annual decrease of 0.95%). Nationwide, reported cases peaked in 2021 at more than 71,000 and remained above 69,000 through 2023^9^.

During the past decade, machine learning techniques have been widely used in public health research to identify relationships between disease outcomes and environmental variables^10^. Time-series approaches, including autoregressive integrated moving average (ARIMA) and seasonal autoregressive integrated moving average (SARIMA) models, have also been applied to infectious disease forecasting in China^11,12^. A study on brucellosis in Xinjiang found that SARIMA outperformed nonlinear autoregressive neural network (NARNN) models, suggesting that seasonal time-series models perform well when brucellosis shows a clear periodic pattern, although the lack of environmental and policy data limited further explanation^13^. To address this limitation, the SARIMA model with exogenous variables (SARIMAX) allows time-variant environmental factors to be incorporated alongside temporal autocorrelation. SARIMAX has been successfully used to model hemorrhagic fever with renal syndrome in China^12^, and a study on hepatitis C prediction showed that incorporating Baidu search index variables reduced forecasting errors compared with SARIMA alone^14^.

Recently, long short-term memory (LSTM), a recurrent neural network (RNN)-based time-series model, has shown strong forecasting ability, including for COVID-19^15,16^. LSTM retains information from previous time steps, making it suitable for temporally correlated data. Its gated structure addresses the vanishing/exploding gradient problems of traditional RNNs^17,18^. In a tuberculosis study in Liaoning, Yang et al. found that a multivariate multi-step LSTM model reduced four error measures by 12.92–14.81% compared with ARIMA^19^.

For brucellosis, existing prediction studies have mainly relied on traditional time-series or neural network models using historical incidence data alone. The performance of LSTM-based models for forecasting brucellosis in high-risk areas of China has not been systematically explored, nor has the added value of LSTM with exogenous variables (LSTMX), which incorporates screened multi-source environmental predictors. This gap is significant because brucellosis transmission is closely linked to livestock exposure, seasonal human activity, ecological conditions, and environmental variation. Therefore, this study aims to evaluate four time-series approaches (SARIMA, SARIMAX, LSTM, and LSTMX) for capturing temporal variability of brucellosis in five highest-incidence provinces (Inner Mongolia, Xinjiang, Shanxi, Heilongjiang, and Hebei) using data from 2014 to 2024, and to identify region-specific forecasting models to inform targeted prevention and control strategies.

## 2. Materials and methods

### 2.1 Data of brucellosis

The time series of the monthly incidence of brucellosis in the top five provinces with the highest national brucellosis incidence in China, including Inner Mongolia Autonomous Region, Xinjiang Uygur Autonomous Region, Shanxi Province, Heilongjiang Province and Hebei Province, from January 2014 to December 2024 was extracted from the China Information System for Disease Control and Prevention. All cases were diagnosed according to the criteria established by the National Health Commission of China. There were no missing values in this data set.

### 2.2 Meteorological data

All monthly meteorological and environmental datasets (2014–2024) for the five studied provinces were collected from authoritative databases. Mean temperature, maximum temperature, minimum temperature, precipitation, and Fractional Vegetation Cover (FVC) were obtained from the National Tibetan Plateau Data Center. Solar radiation, soil moisture, and wind speed were derived from the TerraClimate dataset. Normalized Difference Vegetation Index (NDVI) was acquired from the NASA MOD13A3 dataset. PM_2.5_, PM_10_, ozone, nitrogen dioxide, carbon monoxide, and sulfur dioxide were extracted from the ChinaHighAirPollutants (CHAP) dataset. Relative humidity and atmospheric pressure were obtained from a high-resolution meteorological forcing dataset published in *Scientific Data*. Detailed sources are provided in Table S1.

### 2.3 Variable Selection

A three-step procedure was established to screen exogenous meteorological variables. First, cross-correlation function (CCF) analysis with 12-order seasonal differencing identified optimal lagged relationships between each meteorological factor and monthly brucellosis incidence; variables with statistically significant positive lags were retained, and the lag period with the strongest correlation was selected. Second, variance inflation factor (VIF) testing was applied to the lagged variables with a threshold of VIF < 5, and an iterative stepwise elimination approach sequentially removed variables with excessive VIF values and the weakest correlation with brucellosis incidence. Third, forward stepwise regression with the Akaike Information Criterion (AIC) was employed on the VIF-retained candidate set, using log-transformed monthly brucellosis incidence as the dependent variable; the variable combination that minimized AIC and achieved optimal model fitting was reserved as the final exogenous predictors for multivariate models.

### 2.4 Modeling of SARIMA and SARIMAX

The ARIMA model is a widely applied time-series forecasting method with favorable prediction accuracy^20^. The SARIMA model is an extended version of the ARIMA model that takes seasonal fluctuations and seasonal trends into account. By incorporating seasonal autoregressive (AR) terms, differencing terms, and moving average (MA) terms, the SARIMA model can accurately forecast time-series data under the assumption of stable temporal trends^21^.

The SARIMA model is denoted as SARIMA(p,d,q)(P,D,Q)s, where p, d, q are non-seasonal orders (autoregressive, differencing, moving average), P, D, Q are seasonal orders, and s is the seasonal period (s=12 in this study). The ARIMAX model integrates exogenous variables to improve forecasting performance compared with conventional ARIMA^22^. In this study, we constructed SARIMAX models based on the SARIMA framework to quantify associations between meteorological factors and brucellosis incidence. By introducing exogenous regressors, these models capture inherent seasonal fluctuations while quantifying the driving effects of multi-source environmental variables (e.g., atmospheric pressure and precipitation) on disease risk.

### 2.5 Constructing the LSTM model

LSTM is a deep learning-based time-series model that uses three gating mechanisms (input, forget, and output gates) to capture long-term dependencies while avoiding vanishing/exploding gradients, making it suitable for monthly brucellosis incidence prediction. LSTM and LSTMX models were built using Python and TensorFlow, with LSTMX incorporating lagged meteorological factors identified through CCF, VIF, and stepwise regression in R. Log transformation was applied to incidence data to reduce skewness, and Min-Max normalization scaled all variables to [0,1]. To ensure reproducibility, a fixed random seed was set, GPU disabled, and TensorFlow deterministic mode activated. The last 12 months of data per province served as the independent test set, with the remaining data for training, and a 3-fold time-series cross-validation (chronological order) was used for hyperparameter optimization. Input samples were constructed using a sliding window, and a grid search optimized window length, number of LSTM neurons, and feature combinations, with the optimal model for each province selected based on minimum Root Mean Square Error under cross-validation. A single-layer LSTM architecture (input layer, LSTM hidden layer, Dropout, fully connected output layer) was adopted, using the Adam optimizer with MSE loss, while an early stopping mechanism monitored training loss and restored optimal weights, which together with Dropout mitigated overfitting and improved generalization.

### 2.6 Evaluation and comparison of models

Four evaluation metrics widely used in public health time-series forecasting—Root Mean Square Error (RMSE), Mean Absolute Error (MAE), Mean Absolute Percentage Error (MAPE), and coefficient of determination (R^2^)—were adopted to quantify model performance on both training and test datasets. Smaller RMSE, MAE, and MAPE values indicate higher prediction accuracy, while R² closer to 1 reflects stronger explanatory power for data variation and better predictive performance.

### 2.7 Data analysis

In this study, R version 3.4.1 was used for variable selection and the development of SARIMA and SARIMAX models. Python was utilized to construct the LSTM and LSTMX models. The statistical significance level was set at α = 0.05.

## 3. Results

### 3.1 Statistical description

This study analyzed the five highest-incidence provinces in China from 2014 to 2024: Inner Mongolia (144,908 cases), Xinjiang (63,229), Shanxi (52,832), Heilongjiang (45,238), and Hebei (44,898). These five provinces accounted for the vast majority of national cases, representing the core endemic areas. Annual average cases varied significantly: Inner Mongolia (1097.79±556.23), Xinjiang (479.01±288.28), Shanxi (400.24±205.75), Heilongjiang (342.71±156.25), and Hebei (340.14±159.97). The data showed positive-skewed distributions with discrete high-incidence years.

Temporal trends are shown in Figure 1 and Figures S1–S5. All provinces exhibited prominent seasonal patterns, with annual peaks between April and July and troughs from November to February. Inter-provincial heterogeneity was evident: Inner Mongolia maintained the highest incidence with persistent seasonal oscillations but no clear monotonic trend; Xinjiang showed moderate fluctuations on a stable baseline; Shanxi displayed an initially high incidence followed by gradual decline; Heilongjiang exhibited small but consistent seasonal fluctuations; and Hebei had the lowest incidence with mild, stable seasonal variations.

**Figure 1.**
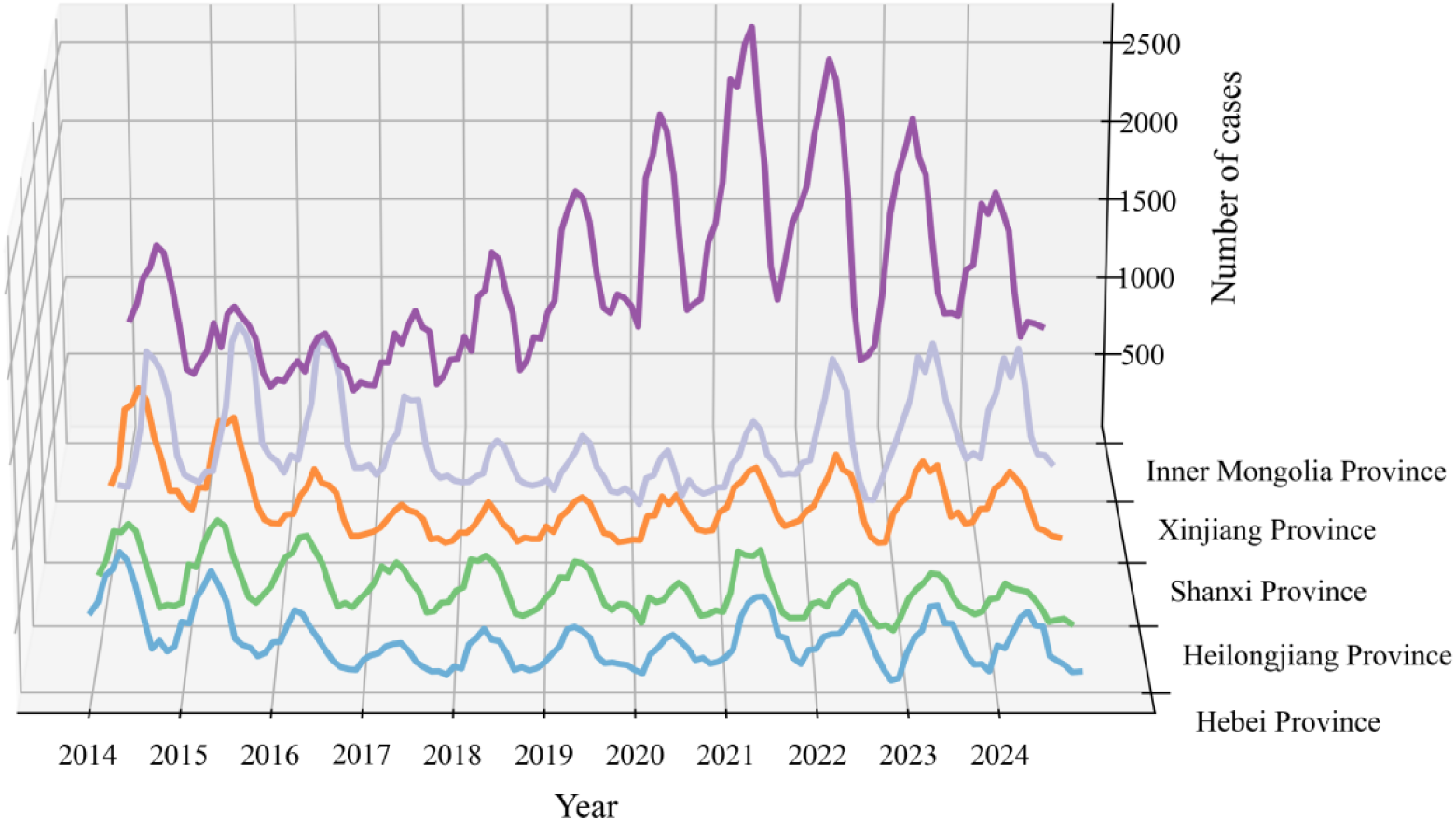
Annual trends of brucellosis cases in five provinces of China, 2014-2024.

Meteorological variables are summarized in Table S2. CCF lag correlation analysis revealed that multiple meteorological factors had significant time-lag correlations with brucellosis cases, with lag effects varying by province. This regional heterogeneity underscores the need to consider province-specific predictors and lag effects when constructing prediction models to improve accuracy and applicability.

### 3.2 Results of Variable Screening

This study applied CCF lag correlation analysis, VIF multicollinearity diagnostics, and AIC stepwise regression to screen environmental factors with significant correlation, no multicollinearity, and optimal AIC values. Cross-correlation heatmaps (Figure S6-S10) visually present the temporal correlation patterns across the five provinces, and Table S3 summarizes the maximum correlation coefficients, optimal lag orders, and VIF values. The correlation magnitude and lag patterns varied substantially across regions.

After systematic screening, the final exogenous covariates were determined: for Inner Mongolia, soil moisture at lag 4; for Xinjiang, ozone at lag 6, soil moisture at lag 1, NO_2_ at lag 1, and precipitation at lag 2; for Shanxi, ozone at lag 6 and wind velocity at lag 1; for Heilongjiang, NDVI at lag 2, PM_10_ at lag 6, and wind velocity at lag 6; for Hebei, ozone at lag 5, SO_2_ at lag 1, and wind velocity at lag 4. These lagged variables were used as exogenous inputs for SARIMAX and LSTMX models.

Figure S10-S15 display the time series of monthly brucellosis cases and selected lagged indicators (standardized by z-score). Distinct regional relationships were observed: in Inner Mongolia, cases were positively correlated with lagged soil moisture; in Xinjiang, negatively correlated with ozone at lag 6 and positively with soil moisture, NO_2_, and precipitation; in Shanxi, ozone showed negative lagged associations while wind velocity showed positive correlation; in Heilongjiang, NDVI, PM_10_, and wind velocity all showed positive lagged trends; in Hebei, lagged ozone was negatively correlated while SO_2_ and wind velocity showed positive correlations. These temporal trends were consistent with the CCF correlation coefficients.

### 3.3 Performance of SARIMA and SARIMAX Models in Predicting Brucellosis Incidence

Various SARIMA and SARIMAX models were established for brucellosis forecasting in five provinces. All models adopted a 12-month seasonal cycle. Performance metrics (AIC, R^2^, RMSE, MAE, MAPE) during the fitting and forecasting stages are presented in Table 1, and the optimal model structures are presented in Table S4.

**Table 1.**
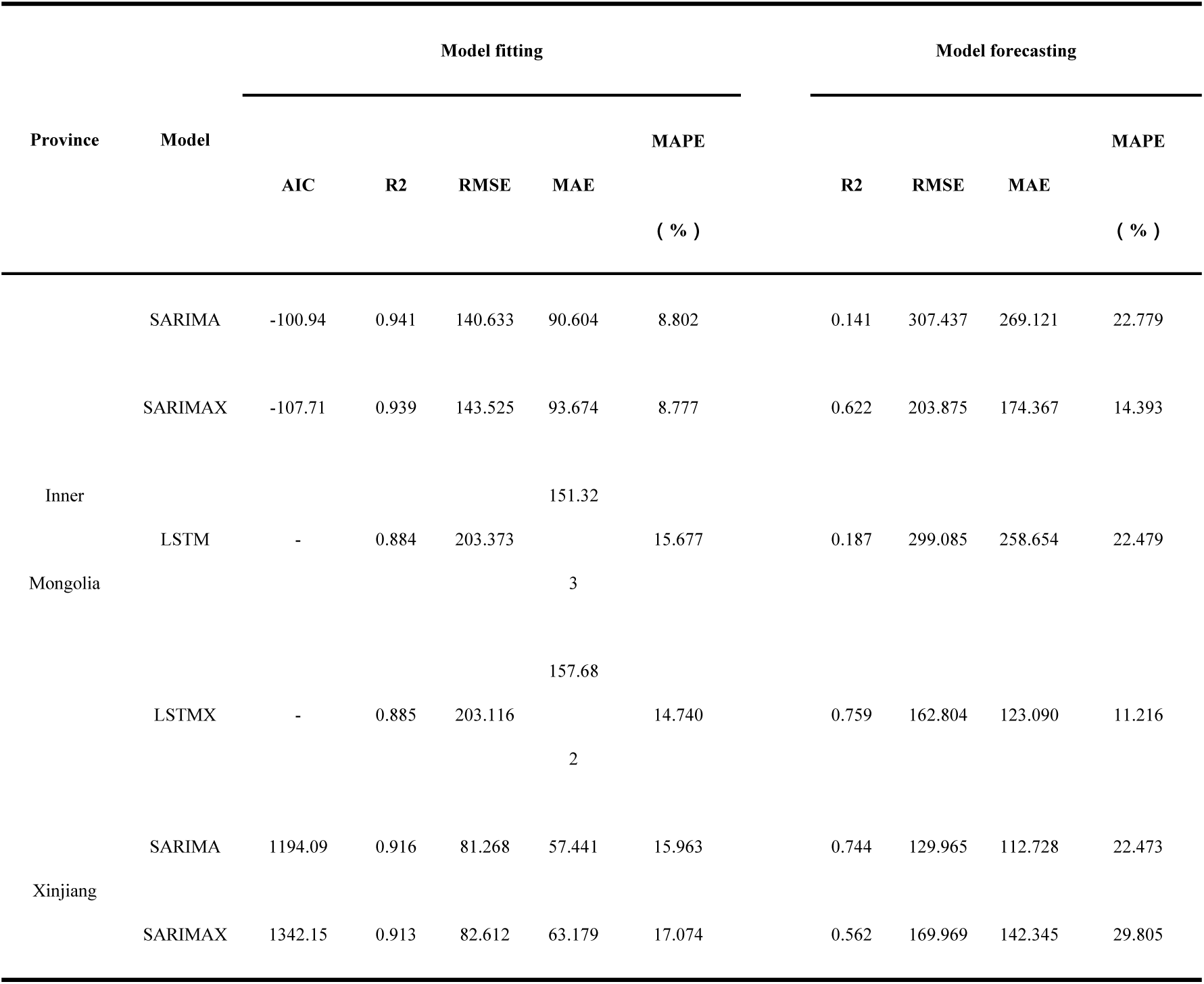
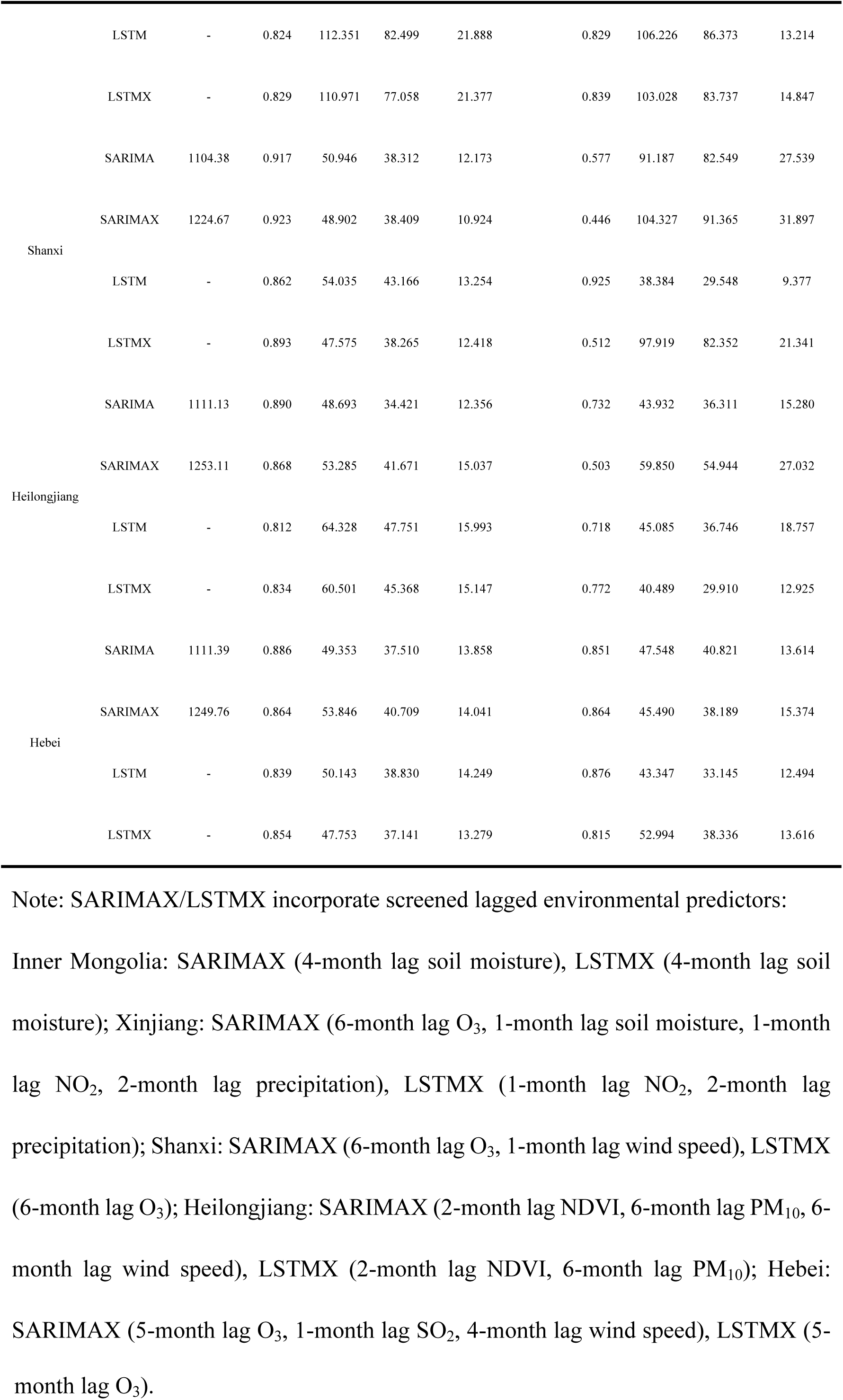
Fitting and prediction performance metrics of SARIMA, SARIMAX, LSTM, and LSTMX for brucellosis in five provinces of China.

At the forecasting stage, model performance varied significantly across provinces. In Inner Mongolia, SARIMAX showed better accuracy than SARIMA, with R^2^ increasing from 0.141 to 0.622, and RMSE, MAE, and MAPE decreasing from 307.437 to 203.875, 269.121 to 174.367, and 22.779% to 14.393%, respectively. In contrast, SARIMAX performed worse than SARIMA in Xinjiang, Shanxi, and Heilongjiang. In Xinjiang, R^2^ decreased from 0.744 to 0.562, while RMSE, MAE, and MAPE increased. In Shanxi and Heilongjiang, SARIMAX also showed lower R^2^ (0.446 vs. 0.577; 0.503 vs. 0.732) and higher errors. For Hebei, SARIMAX showed marginal improvement in R^2^ (0.864 vs. 0.851) and RMSE (45.490 vs. 47.548), but slightly higher MAE (38.189 vs. 40.821) and MAPE (15.374% vs. 13.614%).

### 3.4 Prediction Performance of LSTM and LSTMX Models for Brucellosis Incidence

A global grid search optimized the sliding window length, number of LSTM hidden layer neurons, and meteorological feature combinations for each province based on minimum RMSE under cross-validation. Performance metrics (R², RMSE, MAE, MAPE) for LSTM and LSTMX during fitting and forecasting are summarized in Table 1.

Notable variations in exogenous variables were identified across provinces. For Inner Mongolia, LSTMX incorporated soil moisture (lag 4). In Xinjiang, it included NO_2_ (lag 1) and precipitation (lag 2). Shanxi’s LSTMX used O_3_ (lag 6). Heilongjiang’s LSTMX integrated NDVI (lag 2) and PM_10_ (lag 6). Hebei’s LSTMX adopted O_3_ (lag 5). Optimal sliding window length, neuron number, and batch size also differed by province, reflecting regional variations in brucellosis patterns and heterogeneous meteorological effects.

Marked provincial differences were observed in forecasting performance. In Shanxi, LSTM achieved the best performance among all models (R^2^=0.925), significantly higher than SARIMA (R^2^=0.577) and SARIMAX (R^2^=0.446), with the lowest errors (RMSE=38.384, MAE=29.548, MAPE=9.377%). In Hebei, LSTM (R^2^=0.876) also outperformed SARIMA (R^2^=0.851) and SARIMAX (R^2^=0.864). In Xinjiang, both LSTM (R^2^=0.829) and LSTMX (R^2^=0.839) outperformed SARIMAX (R^2^=0.562). In Inner Mongolia, LSTMX performed best (R^2^=0.759), higher than SARIMA (R^2^=0.141) and SARIMAX (R^2^=0.622), with the lowest errors (RMSE=162.804, MAE=123.090, MAPE=11.216%). Conversely, LSTMX underperformed relative to LSTM in Shanxi (R^2^ dropped from 0.925 to 0.512) and Hebei (R^2^ dropped from 0.876 to 0.815). In Heilongjiang, LSTMX slightly outperformed LSTM (R²: 0.772 vs. 0.718), with lower errors (RMSE: 40.489 vs. 45.085; MAE: 29.910 vs. 36.746; MAPE: 12.925% vs. 18.757%).

### 3.5 The Comparisons of SARIMA,SARIMAX,LSTM and LSTMX Estimations

As shown in Figure 2, the fitting and prediction results are separated by a vertical dashed line. The grey, orange, red, green, and purple lines represent actual cases, SARIMA, SARIMAX, LSTM, and LSTMX estimates, respectively. Table S5 presents the monthly predicted values for each province.

**Figure 2.**
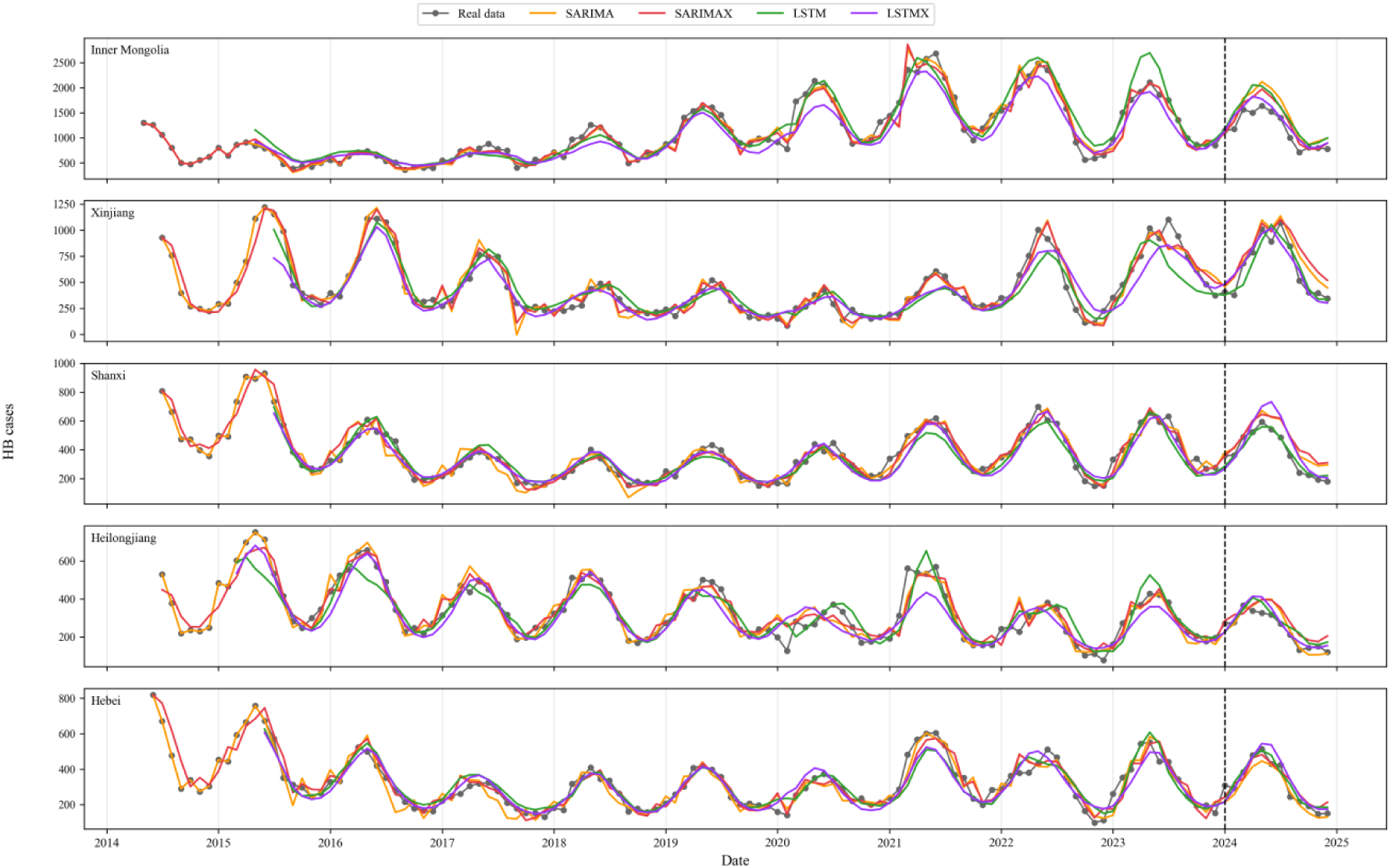
Temporal fitting and prediction curves of four time series models for brucellosis notifications in five provinces of China.

The main findings are as follows: (1) LSTM and LSTMX use a fixed 12-month sliding window, so their fitting curves start later (around 2015) than SARIMA/SARIMAX (2014). (2) LSTM and LSTMX produce smoother estimates with smaller local fluctuations and better capture long-term seasonal trends, while SARIMA/SARIMAX are more sensitive to short-term monthly fluctuations. All five provinces exhibit a consistent unimodal pattern: cases increase from January to May, peak between May and July, then decline from August to December, with no secondary peak. (3) SARIMA and SARIMAX tend to overestimate peak incidence, whereas LSTM and LSTMX provide more conservative and accurate peak estimates. During the low-incidence period (September–December), all four models capture low-level characteristics well, but SARIMA/SARIMAX show slight under– or overestimation in certain months, while LSTM/LSTMX yield more stable estimates with smaller biases. The multivariate LSTMX generally achieves better fitting accuracy than univariate models (SARIMA and LSTM).

## 4. Discussion

### 4.1 Spatiotemporal distribution of human brucellosis

Brucellosis is a severe zoonotic disease with pronounced seasonal fluctuations and regional clustering, remaining a priority for prevention and control in China^23^. The five highest-incidence provinces—Inner Mongolia, Xinjiang, Shanxi, Heilongjiang, and Hebei—collectively determine the national epidemic trend. Between 2014 and 2024, these five provinces accounted for the vast majority of reported cases nationwide, with Inner Mongolia ranking first (annual average: 1097.79 cases). This distribution pattern is consistent with existing studies identifying Inner Mongolia, Shanxi, Heilongjiang, and Hebei as highest-incidence provinces (2005–2010)^24^, with Inner Mongolia and adjacent areas as main high-risk clusters^25^.

All five provinces exhibited a consistent unimodal seasonal pattern, with peak incidence between April and July and a trough in winter. This aligns with brucellosis transmission dynamics: warmer spring/summer temperatures coincide with livestock birthing and increased human outdoor activity, jointly driving epidemic spread. Environmental factors in Xinjiang show both short-lag and cumulative effects (rainfall lag: ∼0.42 months; cumulative effect: ∼1.95 months)^26^. In northeastern China, low temperatures have a longer lag effect (peak lag: 2 months) while high temperatures have a more immediate effect (peak lag: 0.6 months)^27^. Similar seasonality has been observed in Henan, where the annual incidence growth rate reached 34.9% after 2018^28^.

### 4.2 Environmental drivers and region-specific lag effects

This study employed a three-step screening procedure (CCF lag analysis, VIF multicollinearity elimination, and AIC stepwise regression) to identify key predictors for each province, revealing significant regional heterogeneity in driving factors and lag periods. In Inner Mongolia, soil moisture at a lag of 4 months was the only selected exogenous variable, as it indirectly influences Brucella survival in the environment; a meta-analysis confirmed that seroprevalence in dairy cows in traditional northern farming areas exceeds 10%^29^. In Xinjiang, selected variables included O_3_ at lag 6 months (negatively correlated), soil moisture at lag 1 month, NO_2_, and precipitation at lag 2 months, with extreme climatic conditions increasing transmission risk^26^. Results from Shanxi, Hebei, and Heilongjiang further underscored regional specificity, consistent with a Shaanxi study confirming that climatic conditions, livestock farming intensity, and socioeconomic status drive incidence in different subregions^30^.

NDVI and FVC were positively correlated with incidence in most regions, suggesting that higher vegetation cover increases exposure risk. Temperature promoted Brucella survival and reproduction, while precipitation was negatively correlated (possibly due to increased livestock confinement on rainy days). Wind speed was positively correlated, potentially facilitating aerosol spread. Atmospheric pressure, relative humidity, ozone, NO₂, and PM₁₀ also showed significant regional associations, indicating that brucellosis epidemiology results from interactions among multiple meteorological, ecological, and air pollution factors. Optimal lag periods varied between 1 and 6 months across regions, highlighting that early warning models must incorporate region-specific lagged variables to ensure accuracy.

### 4.3 Model performance comparison and applicability analysis

The SARIMA model demonstrated stable fitting performance across all five provinces and effectively captured seasonal fluctuations, consistent with previous brucellosis prediction studies^28,30^. However, its inability to incorporate exogenous variables limits predictive accuracy. The SARIMAX model showed heterogeneous performance across regions: accuracy improved significantly in Inner Mongolia and Hebei but decreased in Xinjiang, Shanxi, and Heilongjiang. This suggests that the benefit of exogenous variables depends on the strength of local environmental drivers. In regions with a single strong driver, such as Inner Mongolia, model performance improved; in regions with complex or weak drivers, incorporating multiple exogenous variables led to overfitting due to limited sample size^31^.

The LSTM model showed clear advantages in most provinces, particularly in Shanxi and Hebei where univariate LSTM achieved the highest prediction accuracy. The LSTMX model performed best in Inner Mongolia and Heilongjiang. This advantage stems from the LSTM gating mechanism, which captures nonlinear dynamics and long-term dependencies in time series data^31,32^. Previous studies have confirmed that LSTM outperforms SARIMA^20^ and that convolutional long short-term memory (ConvLSTM) outperforms traditional machine learning methods^30^. Compared with SARIMA and SARIMAX, LSTM models produce smoother fitted curves, smaller peak prediction deviations, and stronger ability to capture long-term nonlinear patterns, overcoming limitations of linear models^31,32^.

However, introducing exogenous variables did not always improve performance. In Shanxi and Hebei, LSTMX underperformed relative to LSTM^33^. Possible reasons include: (1) environmental factors may have complex interactions or threshold effects not fully captured; (2) correlations between some meteorological variables and brucellosis may be non-causal; (3) multicollinearity screening may not fully address overfitting in time series forecasting^34^. A comparative study on hand, foot, and mouth disease also confirmed that LSTM outperformed SARIMA models^35^.

From a public health perspective, the conservative peak estimation of LSTM models has greater practical value^30^. LSTM and other machine learning methods are more suitable for accurate long-term trend prediction, while SARIMA’s high sensitivity may be more valuable for short-term real-time early warning.

In summary, we propose an adaptive strategy for brucellosis prediction: SARIMA for basic surveillance prioritizing stability and interpretability; LSTM for high-precision early warning; LSTMX or SARIMAX for regions with strong environmental drivers; and avoiding direct cross-regional application of a single model.

### 4.4 Public health and One Health implications for prevention and control

Machine learning methods are widely used for modeling nonlinear systems in public health, providing precise predictive models^36–38^. The seasonal peak characteristics of brucellosis identified in this study provide a clear scientific basis for determining the timing of prevention and control interventions. Given the pronounced spring–summer peak in China, it is recommended to strengthen health education, livestock quarantine, and environmental disinfection before March each year^39^. The region-specific environmental factors identified—soil moisture in Inner Mongolia, ozone and precipitation in Xinjiang, ozone and wind speed in Shanxi, NDVI and PM_10_ in Heilongjiang, and ozone and sulfur dioxide in Hebei—can serve as early warning indicators. Deep learning models such as LSTM can achieve monthly single-step predictions, supporting rational allocation of medical resources and targeted control measures.

Under the One Health framework, human, animal, and environmental health are integrally linked^40^. Brucellosis prevention and control require a comprehensive, cross-sectoral strategy, as animal epidemics are inseparably linked to sustained human transmission^41^. The environmental drivers identified in this study provide a scientific basis for cross-sectoral joint prevention and control. Through multisectoral collaboration among health, livestock, and meteorological authorities, monitoring environmental changes and implementing early intervention measures can effectively reduce the occurrence and spread of brucellosis.

### 4.5 Limitations and future directions

This study has several limitations. First, only 120 months of data were used for training (January 2014 to December 2023) and 12 months for testing (January to December 2024). This limited sample size may have constrained LSTM performance, as insufficient data can lead to overfitting, increased prediction error, and model instability^42^. Second, only environmental factors were incorporated; other important drivers—such as livestock farming scale (e.g., sheep and cattle inventory), immunization and quarantine measures, population mobility, and socioeconomic conditions (e.g., Gross Domestic Product (GDP), per capita disposable income, mutton price and production)—were not included, although these are significantly correlated with brucellosis incidence^43^. Third, this study employed a basic LSTM architecture without exploring hybrid approaches combining attention mechanisms or convolutional neural networks (e.g., convolutional neural network-long short-term memory (CNN-LSTM), ConvLSTM) or SARIMA-LSTM combinations. Existing research shows that CNN-LSTM hybrid models outperform single LSTM models in mitigating overfitting and improving accuracy^44^, ConvLSTM outperforms traditional LSTM^34^, and combined ARIMA–Elman neural networks outperform single ARIMA models^45^.

Future directions include: integrating multisource heterogeneous data (livestock epidemics, socioeconomic activities, ecological data) to build more interpretable and generalizable prediction models; developing hybrid deep learning architectures with attention or convolutional mechanisms to improve accuracy and interpretability; conducting long-term risk projections under different climate change scenarios; and developing a real-time early warning system for dynamic epidemic surveillance and visualization to translate modeling results into practical prevention and control applications.

## 5. Conclusion

In summary, this study systematically compared four time-series models (SARIMA, SARIMAX, LSTM, LSTMX) for brucellosis prediction. LSTM significantly outperformed SARIMA and SARIMAX in most provinces. However, incorporating exogenous meteorological variables did not consistently improve prediction accuracy, and variable selection must balance regional specificity, biological plausibility, and model complexity. Further analysis revealed pronounced regional heterogeneity in the effects of meteorological-environmental factors on brucellosis transmission. These findings support the development of localized early warning systems for brucellosis in high-risk areas of China and provide scientific evidence for understanding region-specific transmission mechanisms.

## Data Availability

The brucellosis incidence data used in this study were obtained from the China Information System for Disease Control and Prevention. These data are owned by a third party and are not publicly available due to confidentiality restrictions. Researchers interested in accessing these data may contact the China Information System for Disease Control and Prevention (https://www.chinacdc.cn) to request permission, subject to the relevant regulations of the National Health Commission of China. All environmental and meteorological data used in this study are publicly available from the sources listed in Table S1 in the Supporting Information, with detailed references and DOIs provided therein. The R and Python code used for model development and analysis are available from the corresponding author upon reasonable request.

## Acknowledgements

We express our gratitude to the China Information System for Disease Control and Prevention for providing the brucellosis incidence data.

